# Pooled testing with replication: a mass testing strategy for the COVID-19 pandemics

**DOI:** 10.1101/2020.04.27.20076422

**Authors:** Julius Žilinskas, Algirdas Lančinskas, Mario R. Guarracino

## Abstract

In absence of a vaccine or antiviral drugs for the COVID-19 pandemic, it becomes urgent to test for positiveness to the virus as many people as possible, in order to detect early outbreaks of the infection. Present testing solutions are based on the extraction of RNA from patients using oropharyngeal (OP) and nasopharyngeal (NP) swabs, and then testing with real-time PCR for the presence of specific RNA filaments identifying the virus. This approach is limited by the availability of reactants, trained technicians and laboratories. To speed up the testing procedures, some attempts have been done on group testing, which means that the swabs of multiple patients are grouped together and tested. Here we propose to use this technique in conjunction with a combinatorial replication scheme in which each patient is allocated in two or more groups to reduce total numbers of tests and to allow testing of even larger numbers of people. Under mild assumptions, a 13× average reduction of tests can be achieved.

## 1 Introduction

The COVID-19 pandemic has already affected more that 2,682,225 people around the world (data retrieved on April 23rd, 2020 from CSSE-JHU repository). It has spread among 212 countries around the world (worldometers.info data).

The fast diffusion of the virus made it clear that in our globalised world threats can spread fast. The lack of knowledge related to the present virus, the contrasting information available, the absence of a targeted vaccine and healing procedures made it soon clear that the diffusion could be mitigated only with restrictions on personal mobility. Quarantine and personal isolation have been applied nearly worldwide. Such restrictions can be made both less stringent and more effective if tests can early detect the presence of infectious individuals. The scarcity of reactants, accredited laboratories for virus testing, and trained technicians have so far severely limited the number of tests.

At the moment of writing, although randomized sampling has not yet been implemented on geographical scale by any country, some attempts are going to be implemented soon to understand the real prevalence and stratification in population by one of the countries that has been first affected by the pandemic outbreak, namely Italy. Such attempts su er from the aforementioned limits in testing capabilities, and therefore only a sample with a size in the order of hundred of thousands individuals can be obtained, with no longitudinal data.

Forty nine counties have executed more than ten thousand tests per million in population. Among those, the prevalence (the number of positive cases among tested individuals) varies between 1 *×* 10*^−^*^5^ and 0.14, with a median of 0.0012. Results are depicted in Fig. 1. This exploratory analysis shows that there are many large countries that would benefit from more efficient testing strategies.

**Figure 1:**
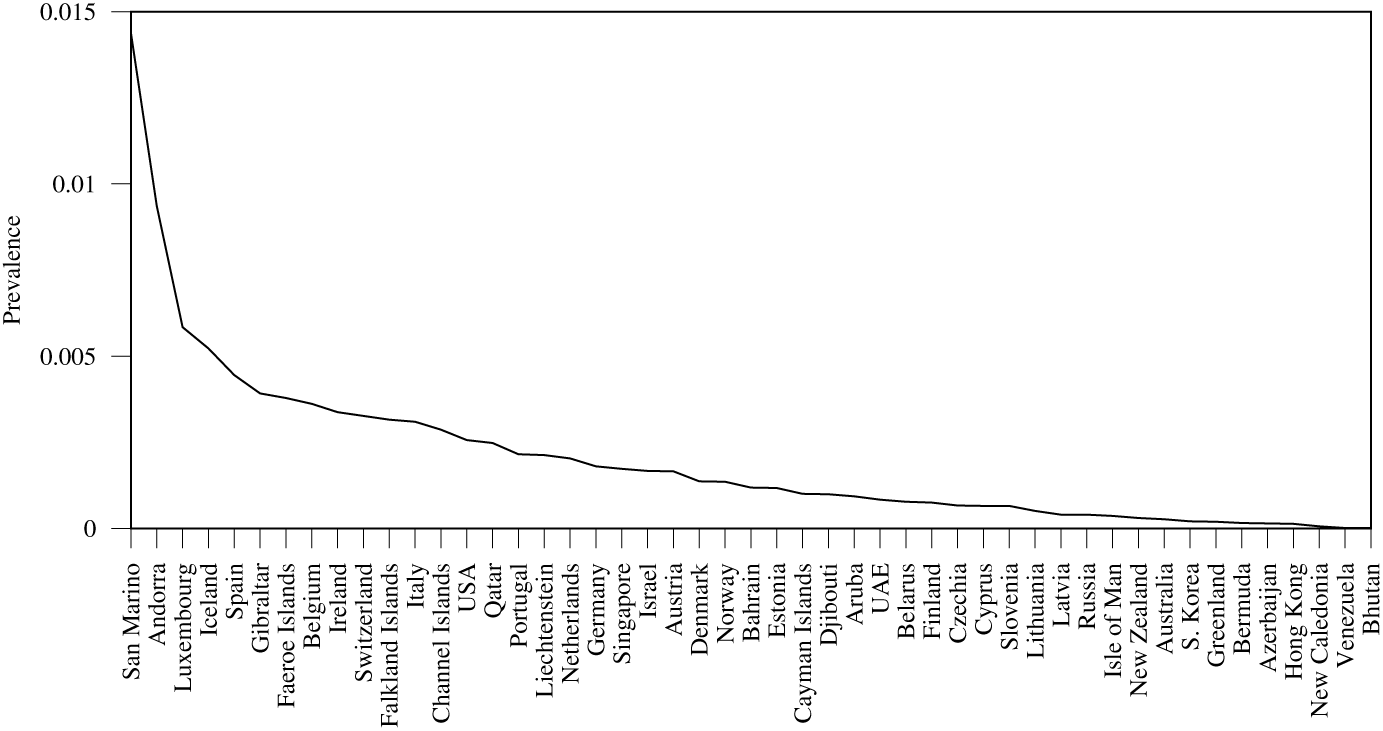
Polygon of the prevalence of the countries with more than ten thousand tests per million population, as April 23rd, 2020 (graphical elaboration of data from worldofmeters.info)

The available tests are based on oropharyngeal and nasopharyngeal swabs and the collected organic material. The virus is then inactivated, and RNA is extracted and amplified. Real-time polymerase chain reaction (RT-PCR) is then used to detect and evaluate the abundance of proteins that are virus-specific. In order to test as many people as possible, studies have shown the possibility of group testing [1], which means that the result of the test is given for a group of people, rather than for a single patient. In the simplest case, this might be applicable to people living in the same apartment, or sharing the same working spaces. In case of a positive person in a group, each individual in the group is singularly tested. It is easy to show that, at low levels of prevalence, such methodologies can provide a decrease in the overall number of tests needed, when compared with individual testing. In some studies, the possibility of using replicas is also analyzed [2]. Pool replicas organize patients in such a way that when one patient is positive, it is possible to directly identify him or her from the positive pools. In case of high prevalence in population, multiple positive patients might appear in the same pools, and other individual tests are necessary to identify them.

Nevertheless, allocation strategies are not optimal with respect to the capability to detect single positive individual, and thus use unnecessary tests. Therefore, we propose an optimized allocation of patients in duplicated pools in order to decrease the total number of tests and the time needed to obtain the results. Such a strategy has already shown [3], in a different context, to decrease the time and cost of diagnosis. We developed an automatic tool to allocate patients to replicated groups that, under certain assumptions, directly provide the information on the positive person in a group [4]. In the present paper, we provide experimental evaluation and comparison of this optimal technique and show, that under reasonable assumptions about prevalence, our proposed methodology always outperforms existing solutions.

The remaining of this work is organized as follow. In Section 2 we review the existing literature. Then, in Section 3 we describe the procedure methodology. Numerical experiments and testing are detailed in Section 4. Conclusions can be found in Section 5.

## 2 Literature review

Experiments indicate that a single positive sample of COVID-19 can be detected in pools of up to 32 samples, with an estimated false negative rate of 10%, and detection of positive samples diluted in even up to 64 samples may also be attainable, though may require additional amplification cycles [5].

This enables pool testing for COVID-19 where the swabs of multiple patients are grouped together and tested. If the result of a sample pool is negative, all the samples in the group are negative. In the case of a positive pool result, individual testing is carried out from previously reserved samples. Depending on the prevalence of disease in population, only a fraction of the groups will need to be tested again for individual results, and therefore the number of tests is reduced accordingly.

For example, suppose 96 samples should be tested and pools of up to 12 samples are possible. In individual testing, 96 tests are necessary. In pool testing, 8 pools of 12 samples are composed and testing is performed. Imagine, that the result of one pool is positive so additional 12 individual tests are needed. Therefore, 20 tests are performed instead of 96. So the number of tests is decreased approximately 5 times with the same number of samples tested. Approximately 5 times more samples could be tested with the same resources. The procedure becomes less efficient when there could be more than one positive sample, e.g. in case of 3 positive samples in different pools, 8 + 12 *×* 3 = 44 tests would be required.

Group testing can be optimised to multiply the power of tests against COVID-19 [6]. For a prevalence of 2%, groups of size 80 are optimal from the statistical point of view. In practice, technical limitations as well as the cost of collection of individual samples put a downwards pressure on group size.

Returning to our example, if 12 pools of 8 samples were used and there were 3 positive samples in different pools, 12 + 8 *×* 3 = 36 tests are required.

Therefore, pool size could be optimized depending on the prevalence. A smaller prevalence enables the use of larger pools.

Other authors [1] propose an adaptive strategy, in which patients with symptoms are individually tested, and pooling is applied only to the asymptomatic ones, in order to reduce the number of positive pools needing individual testing.

In our previous work [3] we proposed optimization of pooled experiments of Next Generation Sequencing for detection of rare mutations. In the study the main aim was to reduce costs and this was achieved by replication of patients in the pools. The proposed replication strategies included transposition and OptReplica. A web-oriented software for the optimization of pooled experiments [4] may be used for optimal planning of pools and replications. A similar idea for replication of patients may be performed in COVID-19 testing but here limited resources are more important than the price.

Group testing for COVID-19 is evaluated in [2] where the testing procedure exactly corresponds to pool testing with transposition based replication [3]. When most tests are negative, pooling reduces the total number of tests up to four-fold at 2% prevalence and eight-fold at 0.5% prevalence.

## 3 Procedure methodology

Replication of patients into pools is performed in such a way that it is possible to directly identify a single positive patient from the positive pools. Let’s illustrate transposition based replication using the example of 96 patients and available pooling of 12 samples. When transposition replication is used, apart from 8 main pools, 12 control/replicated pools are formed with different compositions of samples. One can imagine 8×12 array of samples and main pools may correspond to rows of samples and control pools then correspond to column of samples. This is illustrated in Figure 2. By performing 20 tests (8 main pools and 12 control pools) a single positive sample may be indicated by the positive result of one specific main and one control pools. So the same 20 number of tests, the same resources as using simple pooling, but shorter time because of unnecessary two-step procedure. In the case of 3 positive samples in different pools, additional individual tests will be necessary with the maximal total amount of tests being 29 compared to 44 or 36 tests of simple pooling illustrated in the previous section.

**Figure 2:**
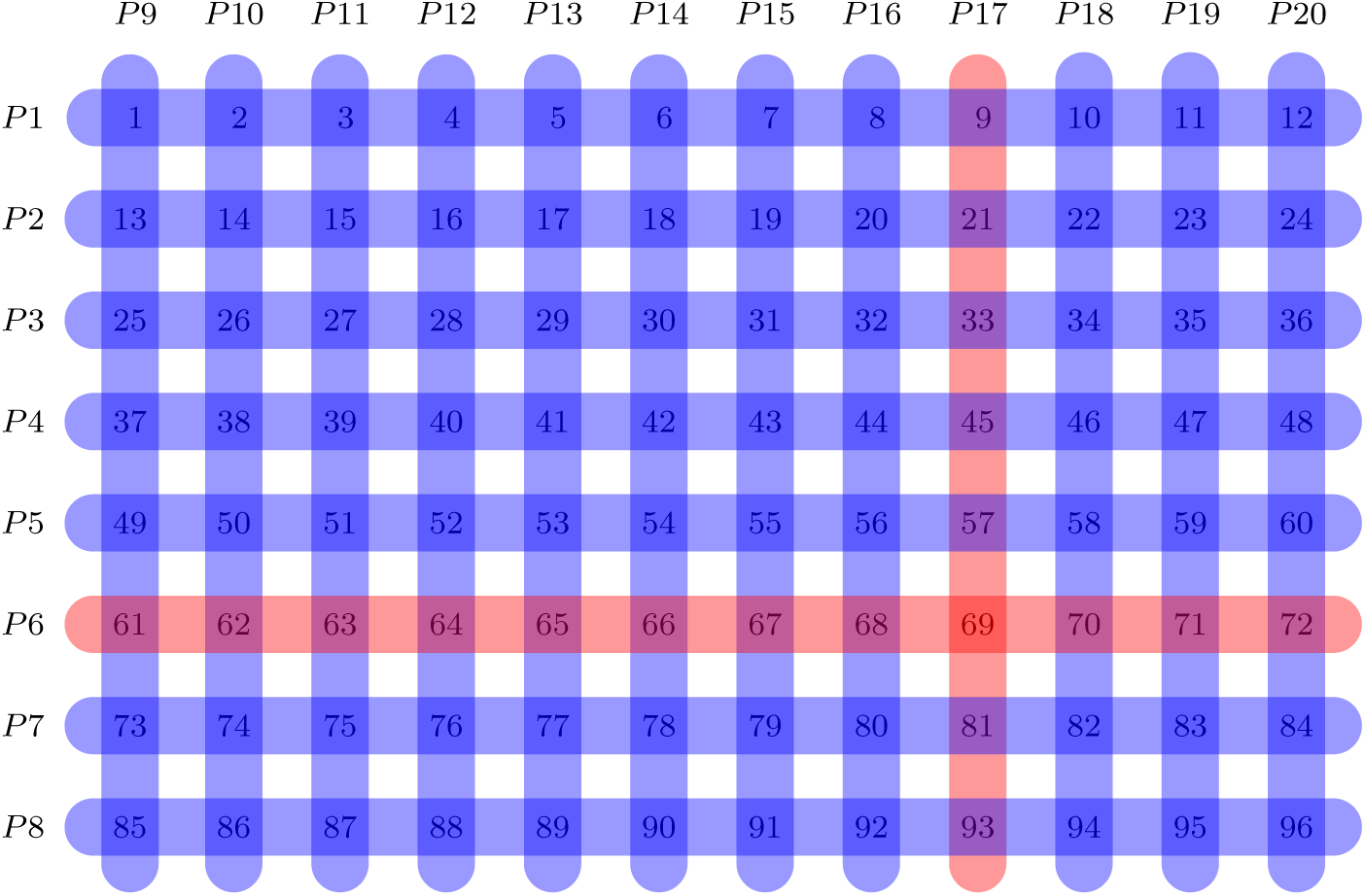
Transposition based replication

Transposition based replication is not the best option when the size and number of pools is different and therefore it is limited when the optimal size of pool is searched. Our proposed OptReplica strategy enables better grouping of patients into pools. Each patient is allocated in the first pool that is not yet completely filled, and replicated in the first pool with the smallest number of allocated patients. Such an allocation of patients guarantees that patient and its replica will be in different pools and logical conjunction of two different pools is the only sample.

For example, 96 patients may be allocated to 16 pools with 12 samples per each and therefore requiring 16 tests to indicate one positive sample out of 96. The allocation and indication are illustrated in Fig. 3. The number of 16 tests is smaller than 20 tests in simple pooling or the same 20 tests in transposition based replication.

**Figure 3:**
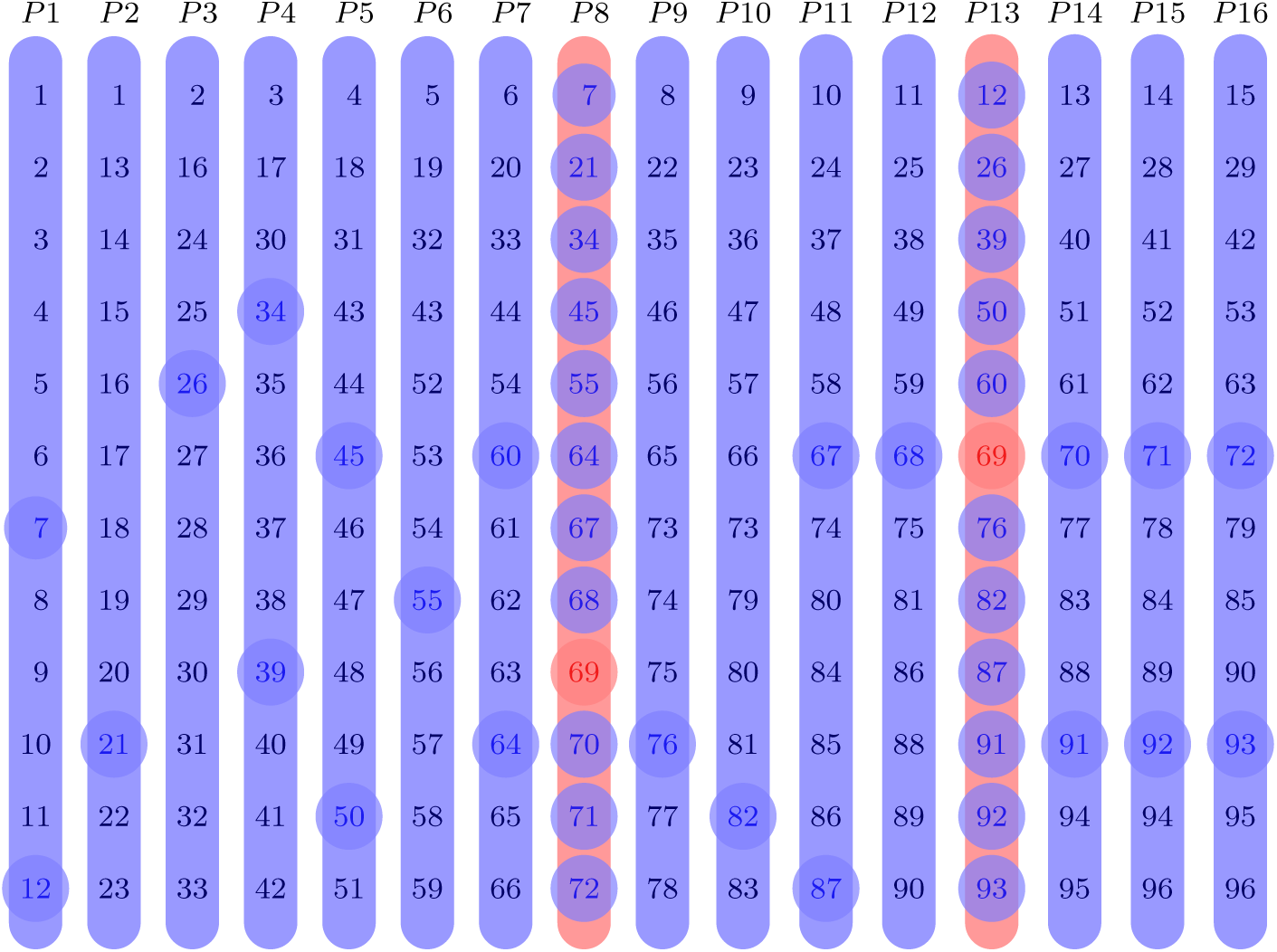
OptReplica replication

After pooling of samples with OptReplica, all pools are tested. If two pools are positive and the remaining pools are negative, then the sample conjugating the positive pools is positive, and the remaining samples in that two pools are negative together with all samples in negative pools.

If more than two pools are positive, then all patients, which samples and their replicas belong to these positive pools must be considered as suspects and individual tests must be performed to validate or negotiate the disease. The number of individual tests can be reduced by analyzing conjunctions of the positive pools; e.g., if we have four positive pools and they are conjugated in pairs, then those two conjugating samples are definitely positive.

In this paper we will focus on the worst case analysis assuming that all patients which samples and their replicas both belong to positive pools must be tested individually, except the situation when only two pools are positive.

For OptReplica to work, the number of pools (*p*) must be larger than the size (*m*) of the largest pool, *p > m*. This implies that the maximal pool size is the largest number *m*, which satisfies the following inequality:

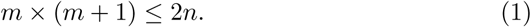

Then the number of pools is:

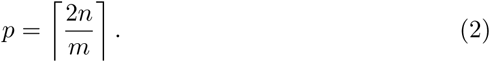

Depending on the probability of positive samples within the target population and the number of possible samples in a pool, optimal allocation of pools may be found. We investigate this in the following section.

## 4 Results and discussion

Efficiency of the proposed strategy OptReplica has been experimentally compared with efficiency of the transposition based replication implemented in accordance with [2].

Two populations of 96 and 384 individuals have been used in order to compare obtained results with those given in [2]. To simulate different prevalence of the disease, each individual of the population is randomly set to be infected with an appropriate probability. For example, in case of prevalence of 1%, the probability to consider an individual as infected is 0.01. Such a random generation of populations simulates different number of infected individuals, which in average correspond to appropriate prevalence, and random distribution of infected among pools. Due to randomness, each experiment has been run for 100 times using randomly generated populations and average results were analysed.

The efficiency of a pooling strategy *S* is expressed by

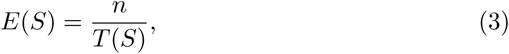

where *n* is the number of samples to be tested and *T* (*S*) is the number of tests performed to identify all infected samples using a pooling strategy *S*. In case of individual testing which requires to test all samples, *E*(*S*) = 1. If a strategy *S* requires two times less tests than the individual testing, then *E*(*S*) = 2. Larger efficiency value means better efficiency of a strategy.

The experimental investigation of transposition based replication using 8 columns by 12 rows pooling of 96 samples and 16 columns by 24 rows pooling of 384 patients gave very similar results to those published in [2].

Several pool sizes (*m ∊ {*4, 6, 8, 12*}*) which divide 2 *×* 96 = 192 samples into completely filled pools and the largest possible pool size (*m* = 13) for replication of 96 samples were selected for OptReplica. Results of all experiments are presented in Table 2. Comparison of efficiency in testing 96 samples using transposition based replication and OptReplica is illustrated in Fig. 4. Here the horizontal axis stands for the prevalence of the disease in percents and the vertical axis – for the efficiency of a pooling strategy. The dashed curve indicates efficiency of transposition based replication and continuous curves indicate efficiency of OptReplica with selected pool sizes.

**Table 1:** Optimal pool sizes for OptReplica with respect to the prevalence of the disease.

| $n$ | Prevalence | | | | | | |
| --- | --- | --- | --- | --- | --- | --- | --- |
|  | 0.1% | 0.2% | 0.5% | 1% | 2% | 5% | 10% |
| 96 | 13 | 13 | 13 | 13 | 13 | 12 | 8 |
| 384 | 27 | 27 | 27 | 27 | 16 | 8 | 6 |

**Figure 4:**
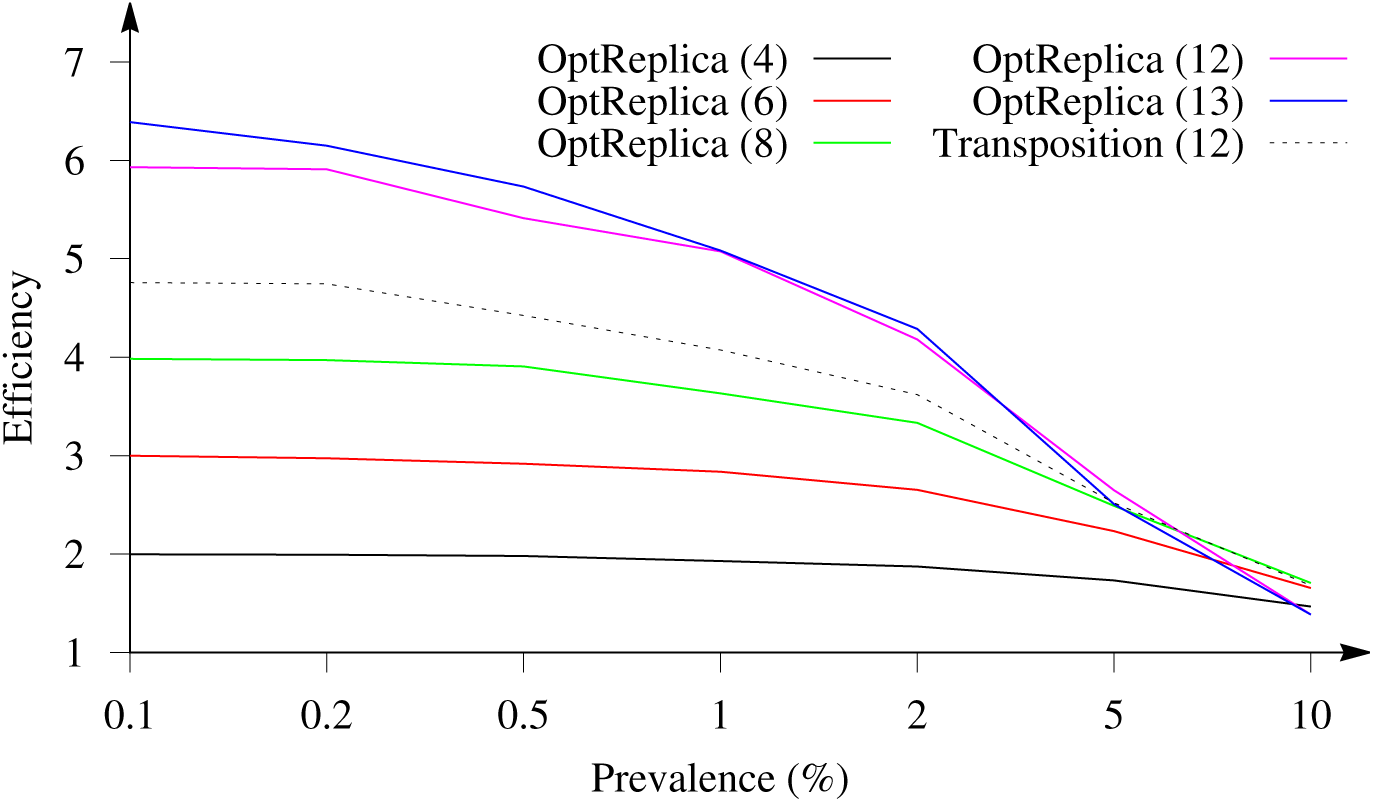
Dependence of efficiency of OptReplica with different pool sizes and transposition based replication on prevalence of the disease testing 96 samples

One can see from the figure, replication strategy OptReplica can reduce by more than 6 times the number of tests needed to determine infected patients in population of 96 individuals, when prevalence is low. In comparison, using the transposition based replication the number of tests can be reduced by less than 5 times by the same prevalence. The best results with OptReplica were achieved using large pools (up to 12 and 13 samples per pool) when the prevalence is lower than 5%. In case of larger prevalence, it is better to use smaller pools (8 samples per pool) for OptReplica.

Similar results have been obtained with *n* = 384 patients. Here, pool sizes (*m ∊* {4, 6, 8, 12, 16, 24}) which divide 2 × 384 = 768 samples into completely filled pools and the largest possible pool size (*m* = 27) for replication of 384 samples were selected for OptReplica. Results of all experiments are presented in Table 3. Selected results are illustrated in Fig. 5, from which we can see that replication strategy OptReplica can reduce by around 13 times the number of tests needed to determine infected patients, when the prevalence is small. Such an efficiency is notably better than the efficiency of the transposition based replication by the same prevalence. Smaller pools must be chosen for OptReplica when the prevalence reaches 1–2 percents. However, even when the prevalence is low, choosing the right pool sizes OptReplica demonstrated notably better efficiency than the transposition based replication by the same prevalence.

**Figure 5:**
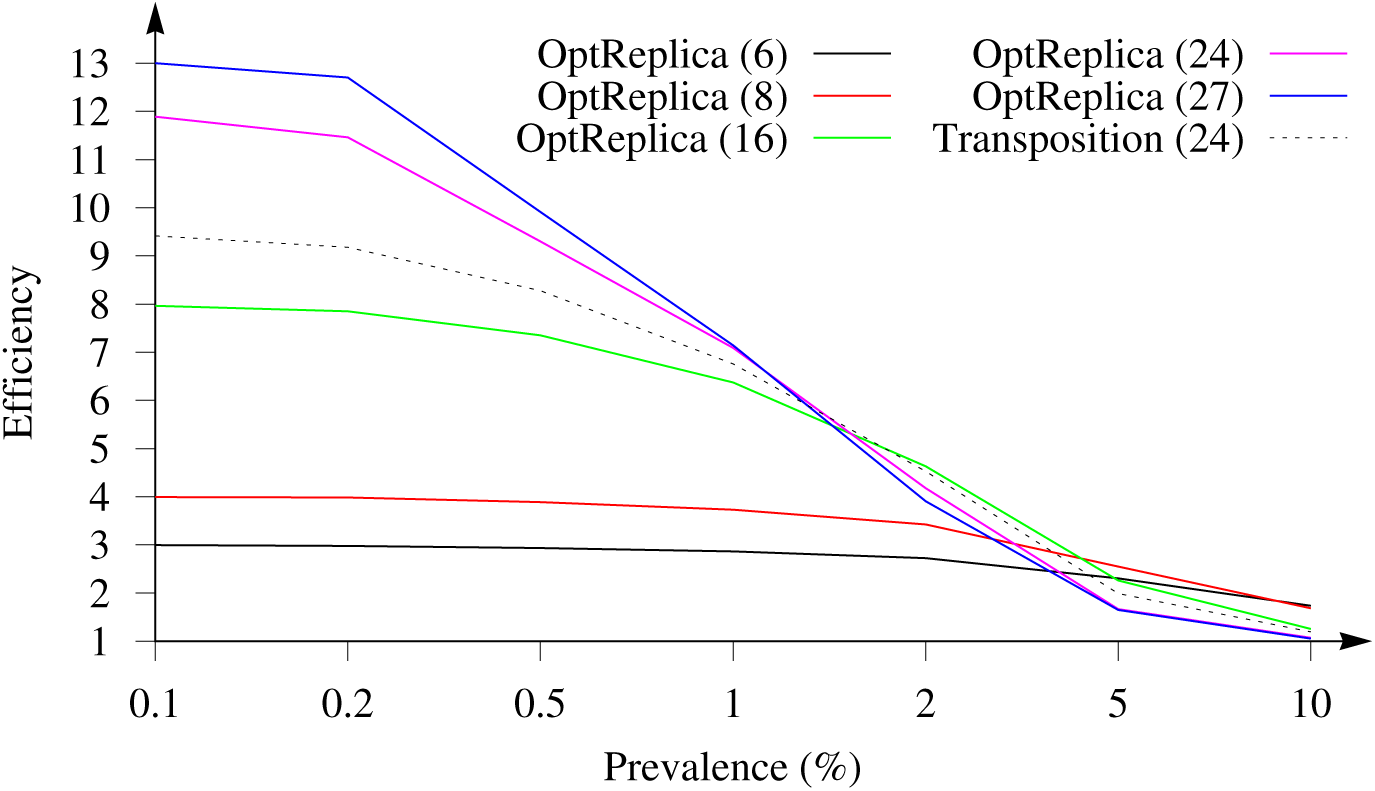
Dependence of efficiency of OptReplica with different pool sizes and transposition based replication on prevalence of the disease testing 384 samples

The obtained results show that independent on the prevalence of the disease OptReplica in average gives better efficiency than the transposition based replication if the right pool size is used: larger pools if the prevalence is smaller and smaller pools, if prevalence is larger. Fig. 6 illustrates comparison of the efficiency of OptReplica with the optimal pool size, chosen depending on the prevalence, with efficiency of the transposition base replication.

**Figure 6:**
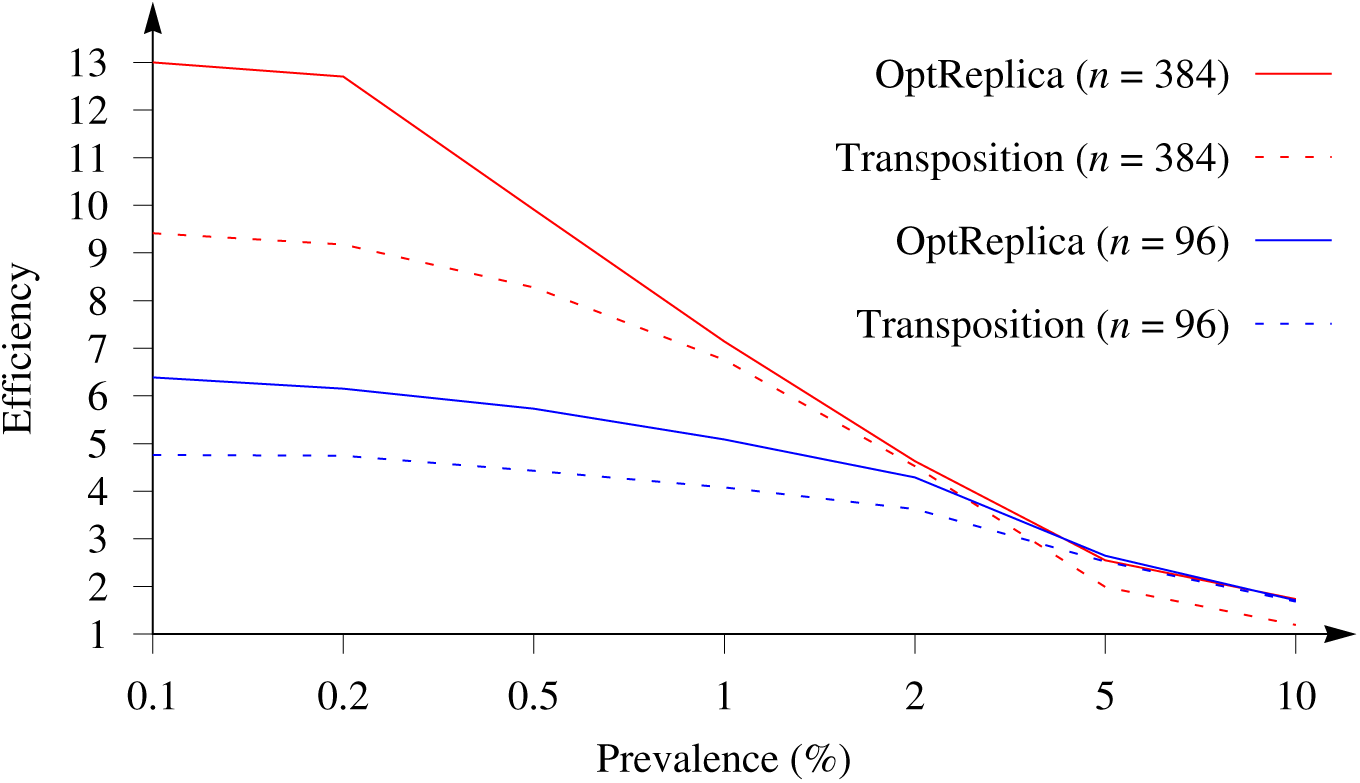
Efficiency of OptReplica with the optimal pools sizes versus transposition based replication

Results of the experimental investigation presented in Figs. 4 and 5 showed that OptReplica can effectively determine positive samples with using more than 13 times less tests, comparing with individual testing when all patients are tested. Although, efficiency of the proposed strategy depends on the prevalence of the disease, choosing the right pool size makes OptReplica better than transposition based replica independent on the prevalence. The optimal pools for testing 96 and 384 with respect to the prevalence of the disease is presented in Table 1.

## 5 Conclusions

In this paper we propose a testing scheme with pools and replica to detect patients positive to COVID-19. We provide examples on how to allocate patients in pools with a strategy that is optimal and minimizes the number of required tests. We show that for different levels of prevalence, it is always possible to find a grouping of the patients that minimizes the number of tests. Our strategy outperforms all other existing techniques, and can reduce the overall number of tests up to 13× times on average.

## Data Availability

All data are available from other studies or websites, i.e. worldometers.info

## 6 Acknowledgments

The work of Mario R. Guarracino was prepared within the framework of the Basic Research Program at the National Research University Higher School of Economics (HSE). The results of Mario R. Guarracino were obtained with the support of the RFFI grant.

### A Appendix

**Table 2:** Efficiency of OptReplica and transposition based replication testing 96 samples. Best results for each prevalence are marked in bold.

| Pooling | $p$ | $m$ | Prevalence | | | | | | |
| --- | --- | --- | --- | --- | --- | --- | --- | --- | --- |
|  |  |  | 0.1% | 0.2% | 0.5% | 1% | 2% | 5% | 10% |
| OptReplica | 48 | 4 | 2.00 | 2.00 | 1.99 | 1.93 | 1.86 | 1.73 | 1.47 |
|  | 32 | 6 | 3.00 | 2.97 | 2.92 | 2.84 | 2.65 | 2.23 | 1.66 |
|  | 24 | 8 | 3.99 | 3.97 | 3.91 | 3.63 | 3.33 | 2.488 | **1.71** |
|  | 16 | 12 | 5.93 | 5.91 | 5.41 | 5.06 | 4.18 | **2.65** | 1.39 |
|  | 15 | 13 | **6.39** | **6.15** | **5.73** | **5.08** | **4.29** | 2.51 | 1.38 |
| Transpos. | 8 | 12 | 4.76 | 4.75 | 4.43 | 4.07 | 3.62 | 2.52 | 1.68 |

**Table 3:** Efficiency of OptReplica and transposition based replication testing 384 samples. Best results for each prevalence are marked in bold.

| Pooling | $p$ | $m$ | Prevalence | | | | | | |
| --- | --- | --- | --- | --- | --- | --- | --- | --- | --- |
|  |  |  | 0.1% | 0.2% | 0.5% | 1% | 2% | 5% | 10% |
| OptReplica | 192 | 4 | 2.00 | 1.99 | 1.98 | 1.95 | 1.90 | 1.74 | 1.50 |
|  | 128 | 6 | 2.99 | 2.98 | 2.93 | 2.86 | 2.73 | 2.30 | **1.74** |
|  | 96 | 8 | 3.99 | 3.98 | 3.88 | 3.73 | 3.42 | **2.55** | 1.68 |
|  | 64 | 12 | 5.97 | 5.95 | 5.59 | 5.17 | 4.32 | 2.53 | 1.47 |
|  | 48 | 16 | 7.96 | 7.85 | 7.35 | 6.37 | **4.63** | 2.26 | 1.26 |
|  | 32 | 24 | 11.89 | 11.46 | 9.30 | 7.09 | 4.17 | 1.66 | 1.07 |
|  | 29 | 27 | **13.00** | **12.70** | **9.91** | **7.14** | 3.90 | 1.65 | 1.05 |
| Transpos. | 16 | 24 | 9.41 | 9.17 | 8.28 | 6.76 | 4.52 | 1.99 | 1.19 |

## References

[1] C. Mentus, M. Romeo, C. DiPaola, Analysis and applications of adaptive group testing methods for COVID-19, medRxiv (2020). doi:10.1101/2020.04.05.20050245.

[2] N. Sinnott-Armstrong, D. Klein, B. Hickey, Evaluation of group testing for SARS-CoV-2 RNA, medRxiv (2020). doi:10.1101/2020.03.27.20043968.

[3] J. Žilinskas, A. Lančinskas, M. R. Guarracino, Application of multi-objective optimization to pooled experiments of next generation sequencing for detection of rare mutations, Plos One 9 (9) (2014) 1–9. doi:10.1371/journal.pone.0104992.

[4] D. Evangelista, A. Zuccaro, A. Lančinskas, J. Žilinskas, M. R. Guarracino, A web-oriented software for the optimization of pooled experiments in NGS for detection of rare mutations, BMC Research Notes 9 (1) (2016) 111. doi:10.1186/s13104-016-1889-6.

[5] I. Yelin, N. Aharony, E. Shaer-Tamar, A. Argoetti, E. Messer, D. Berenbaum, E. Shafran, A. Kuzli, N. Gandali, T. Hashimshony, Y. Mandel-Gutfreund, M. Halberthal, Y. Ge en, M. Szwarcwort-Cohen, R. Kishony, Evaluation of COVID-19 RT-qPCR test in multi-sample pools, medRxiv (2020). doi:10.1101/2020.03.26.20039438.

[6] C. Gollier, O. Gossner, Group testing against Covid-19, Covid Economics 2 (2020) 32–42.

